# Transcranial Focused Ultrasound for Emotion Regulation: A Systematic Review and Quantitative Summary of Human Studies

**DOI:** 10.1101/2025.09.03.25334438

**Authors:** Youbin Kang, Kyu-Man Han, Byung-Joo Ham, Dorothee P. Auer, Marcus Kaiser, JeYoung Jung

## Abstract

**Background:** Emotion regulation is a core transdiagnostic process in mood, anxiety, and stress-related disorders. While existing non-invasive brain stimulation approaches, such as transcranial magnetic stimulation (TMS) and transcranial direct current stimulation (tDCS), can modulate affective networks, their clinical use is limited by restricted spatial precision and depth penetration. Transcranial focused ultrasound stimulation (tFUS) offers submillimeter focality and access to both cortical and deep subcortical structures, making it a promising tool for affective neuromodulation.

**Methods:** We systematically searched PubMed, Embase, and PsycINFO for human studies using tFUS to modulate emotion regulation, affective processing, or related symptoms. Eligible studies included randomized controlled trials (RCTs), open-label, and within-subject designs. Data on stimulation parameters, target regions, outcomes, and safety were extracted. Effect sizes were calculated and pooled using a random-effects model, with subgroup analyses by clinical domain.

**Results:** Eleven studies met inclusion criteria, targeting the amygdala (n = 5), prefrontal cortex (n = 5), or subcallosal cingulate cortex (n = 1), with protocols varying in frequency (250–650 kHz), duty cycle (0.5–70%), and number of sessions (1–25). Across six studies reporting behavioral symptom outcomes, the pooled effect was moderate-to-large (Hedges’ g = 0.88, 95% CI [0.47, 1.29]), with larger effects in depression-related measures (g = 1.31) than in anxiety-related measures (g = 0.67). Imaging outcomes were reported in a smaller subset of studies and were not included in the pooled estimates. No serious adverse events were reported.

**Conclusions:** tFUS is a safe and well-tolerated intervention capable of engaging deep affective circuits. Future large-scale, harmonized, and mechanistically informed trials are warranted to refine protocols, establish durability, and optimize translation into clinical practice.

## Introduction

Emotion regulation is a core psychological function that supports adaptation to internal states and external demands (Gross, 2015). Its dysfunction is a transdiagnostic feature across a wide range of psychiatric conditions (Lincoln et al., 2022), including major depressive disorder (Villalobos et al., 2021), generalized anxiety disorder (Cisler & Olatunji, 2012; Eres et al., 2021), and post-traumatic stress disorder (McLean & Foa, 2017). Consequently, the modulation of emotion regulation networks has emerged as a promising target for both prevention and intervention strategies across mental health disorders (Regenold et al., 2022; Tanaka et al., 2025).

Despite advances in pharmacological and psychotherapeutic treatments, a substantial proportion of patients experience suboptimal responses or relapse, highlighting the need for novel therapeutic approaches (Guidetti et al., 2025; Nierenberg et al., 2003). Non-invasive brain stimulation techniques, including transcranial magnetic stimulation (TMS) and transcranial direct current stimulation (tDCS), have been explored for their capacity to modulate affective neural circuits (Abend et al., 2019; Andò et al., 2021; Bertocci et al., 2021; Liu et al., 2017; Oathes et al., 2021). While these approaches have demonstrated efficacy in selected populations, their clinical impact remains constrained by limited spatial specificity, variable penetration depth, and inconsistent long-term outcomes (Aderinto et al., 2024; Tao et al., 2024; Razza et al., 2023). Recent reviews suggest that most neuromodulation protocols may be effectively “underdosed,” contributing to modest response rates and motivating accelerated treatment schedules (Sabé et al., 2024; Sharif et al., 2025). Safety and tolerability also remain important considerations, with adverse effects reported across the spectrum from serious events such as mania, psychosis, and seizures to more common outcomes such as headaches and tinnitus (Kang et al., 2024; Toth et al., 2024; Zhou et al., 2024).

Transcranial focused ultrasound stimulation (tFUS) has recently emerged as a novel and potentially transformative neuromodulatory technique, attracting attention in the fields of neuroscience and clinical translational research while being a preferred intervention option by patients suffering from brain and mental health conditions (Atkinson-Clement et al., 2025). Utilizing highly focused acoustic energy, tFUS enables superior focal precision, with the capacity to target both cortical and deep subcortical regions while preserving millimeter-scale resolution and safety profile (Badran & Peng, 2024; Jin et al., 2024). Unlike high-intensity focused ultrasound, used for ablative purposes, low-intensity tFUS (LIFU) operates below thermal thresholds, leveraging mechanical and neurochemical mechanisms to modulate neuronal excitability without inducing thermal or structural damage (Fini & Tyler, 2017; Zhang et al., 2021). This unique capability positions tFUS as an ideal candidate for engaging distributed emotion regulation networks, including deep structures such as the amygdala and anterior cingulate cortex. As such, preliminary human studies have demonstrated that low-intensity tFUS can modulate mood states, physiological arousal, and brain network dynamics (Chou et al., 2024; Kim et al., 2022; Sanguinetti et al., 2020). However, the emerging literature remains methodologically heterogeneous, with considerable variability in stimulation parameters, targeted brain regions, outcome measures, and study populations (Caffaratti et al., 2025).

To advance the field and guide future translational applications, the present systematic review aims to comprehensively summarize human studies that have employed tFUS with the explicit intent of modulating emotion-related outcomes. Specifically, we examine patterns across stimulation protocols, identifying convergent and divergent effects on affective endpoints, and evaluate consistency and safety of reported neuromodulatory outcomes. In doing so, we aim to provide a comprehensive overview of current evidence and identify key parameters and methodological considerations that can inform future research and therapeutic development in the field of affective neuromodulation.

## Methods

The protocol for this study was developed in accordance with the Preferred Reporting Items for Systematic Reviews and Meta-Analyses (PRISMA) guidelines (Moher et al., 2009; Page et al., 2021) and has been published on PROSPERO (Registration No. CRD1102146). Studies were identified by searching PubMed/Medline, Embase and PsycINFO using the following search terms (and/or their variants): transcranial focused ultrasound, low-intensity focused ultrasound, ultrasound neuromodulation, depression, anxiety, post-traumatic stress disorder (PTSD), psychiatric disorder, and emotion regulation. Complete database-specific strategies, including field tags, controlled vocabulary (MeSH/Emtree/Thesaurus), synonyms, and limits are provided in Supplement S1. No year limits were imposed a priori. In addition to the database search, we manually screened the reference lists of all included studies and relevant reviews to identify additional articles that may not have been captured in the initial search.

### Inclusion and Exclusion Criteria

Studies were eligible for inclusion if they (1) involved human participants (healthy volunteers or clinical populations), (2) applied transcranial focused ultrasound (tFUS) with neuromodulatory intent (i.e., excluding blood-brain barrier opening studies), and (3) reported outcomes related to emotion regulation, affective processing, or symptoms of anxiety, depression, or stress. We included randomized controlled trials (RCTs), non-randomized trials, and within-subject experimental designs.

Studies were excluded if they (1) were conducted on animals or *in vitro* models, (2) lacked any emotional, affective, or stress-related outcomes, (3) used ultrasound solely for diagnostic imaging or mechanical BBB opening, or (4) were review articles, commentaries, or conference abstracts without primary data.

### Study Selection

All identified records were imported into reference management software (EndNote) for deduplication using exact/loose matching of Title, Author, Year, and DOI. Two reviewers (Y.K. and J.J.) independently screened titles and abstracts for eligibility. Full-text reviews were then conducted for all potentially relevant articles. Discrepancies at either stage were resolved through discussion and consensus. Where eligibility remained unclear, a third reviewer was consulted.

In total, 319 citations were identified. After removing duplicates, 272 unique records were screened. Of these, 24 full-text articles were retrieved for further review, and 11 studies were deemed eligible for inclusion in the qualitative synthesis, with 6 studies included in the quantitative meta-analysis.

The PRISMA 2020 flow diagram (Figure 1) details the selection/elimination process and numbers at each stage.

**Fig 1.**
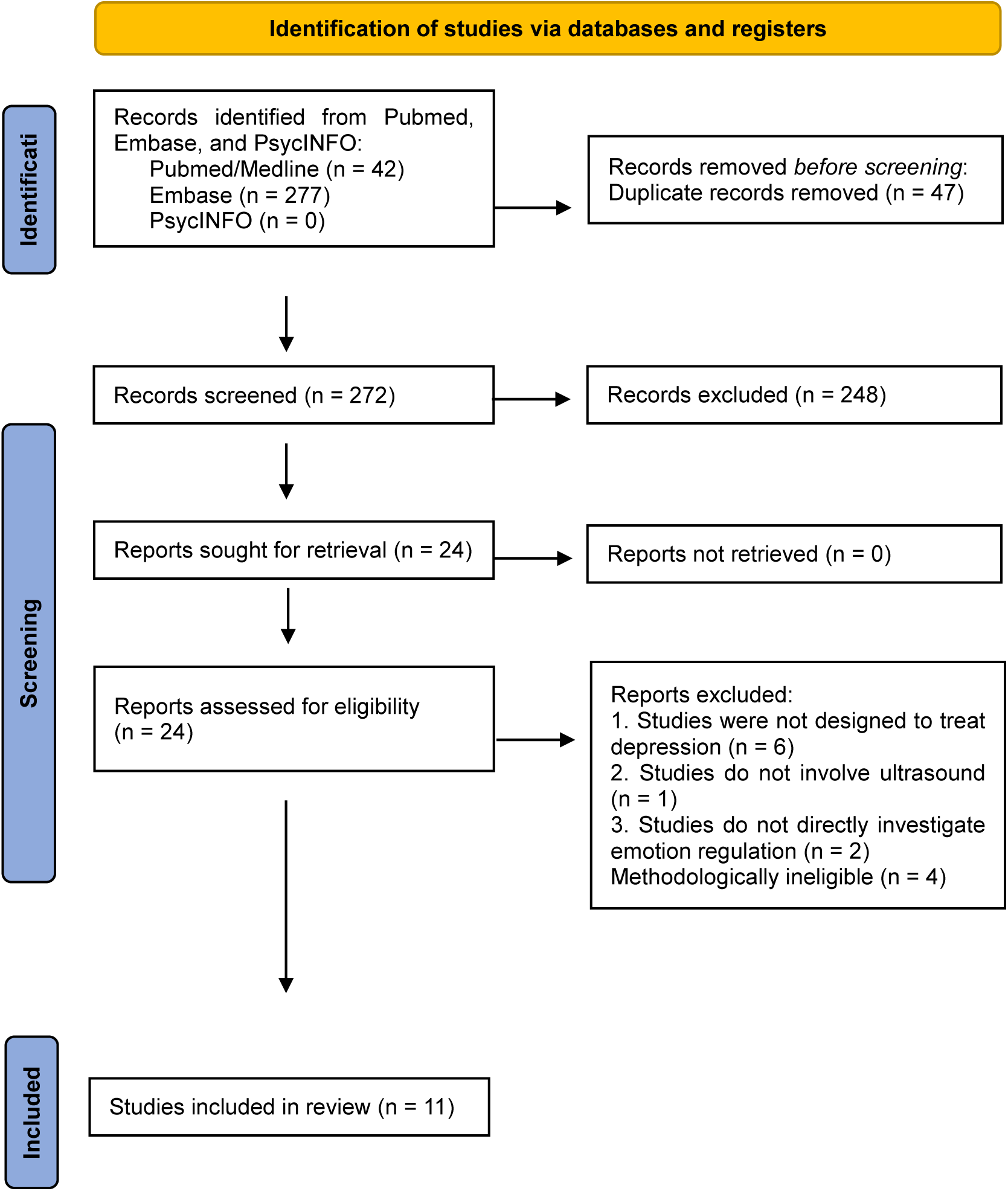
PRISMA flow diagram for study identification and selection.

### Data Extraction

For each included study, the following data were extracted: (1) publication details (author, year, journal), (2) sample characteristics (sample size, age, sex, clinical status), (3) stimulation protocol (target brain region, acoustic intensity at the target region, frequency, pulse duration, number of sessions), (4) study design, (5) outcome measures related to affect, emotion, or mood, (6) control condition (sham, baseline, or active comparator), and (7) reported adverse events or safety concerns.

Where available, pre- and post-intervention scores on validated clinical or behavioral outcome measures were extracted to calculate standardized effect sizes (Hedges’ g). Hedges’ g, a bias-corrected version of Cohen’s d, provides more accurate estimates in small-sample studies (Hedges & Olkin, 2014). 95% confidence intervals were computed under a random-effects model, with small-sample bias correction applied when appropriate.

### Quality and Risk of Bias Assessment

Given the variability in designs across included studies, we adapted key criteria from the Cochrane RoB 2.0 and the NIH Quality Assessment Tools (Supplementary Table 1). Each study was assessed across five domains: (1) clarity of randomization/blinding, (2) adequacy of control condition, (3) validity of outcome measures, (4) completeness of reporting, and (5) consistency of follow-up and analysis procedures. Risk of bias was rated as low, moderate, or high in each domain, with discrepancies resolved through discussion and consensus.

### Quality Synthesis

Where sufficient data was available, a meta-analysis was conducted on a subset of studies that reported validated pre- and post-intervention scores on mood- or anxiety-related measures (e.g., Hamilton Anxiety Rating Scale (Hamilton, 1959), Montgomery-Å sberg Depression Rating Scale (Montgomery & Å sberg, 1979), State-Trait Anxiety Inventory (Spielberger et al., 1971)). Standardized effect sizes were calculated as Hedges’ g based on mean difference and pooled standard deviation, applying a bias correction for small samples. A random-effects model was used to account for expected heterogeneity in stimulation targets and study populations. Subgroup analyses by clinical domain were conducted to evaluate potential moderators of treatment response.

Sensitivity analyses were performed excluding ultra-small samples (N < 10) to assess robustness. Additional exploratory restrictions (e.g., patient-only, blinded only, leave-one-out) were considered but results were not interpretable due to limited number of available studies.

## Results

### Study Characteristics

A total of 11 studies met inclusion criteria, encompassing both healthy and clinical populations diagnosed with conditions such as major depressive disorder (MDD), generalized anxiety disorder (GAD), or post-traumatic stress disorder (PTSD). Sample sizes ranged from 5 to 152, with participants spanning a wide adult age range. Study designs included randomized controlled trials, open-label pilot studies, and within-subject crossover experiments. All studies employed tFUS to modulate emotion-related outcomes using either inhibitory or excitatory protocols (Table 1).

**Table 1.**
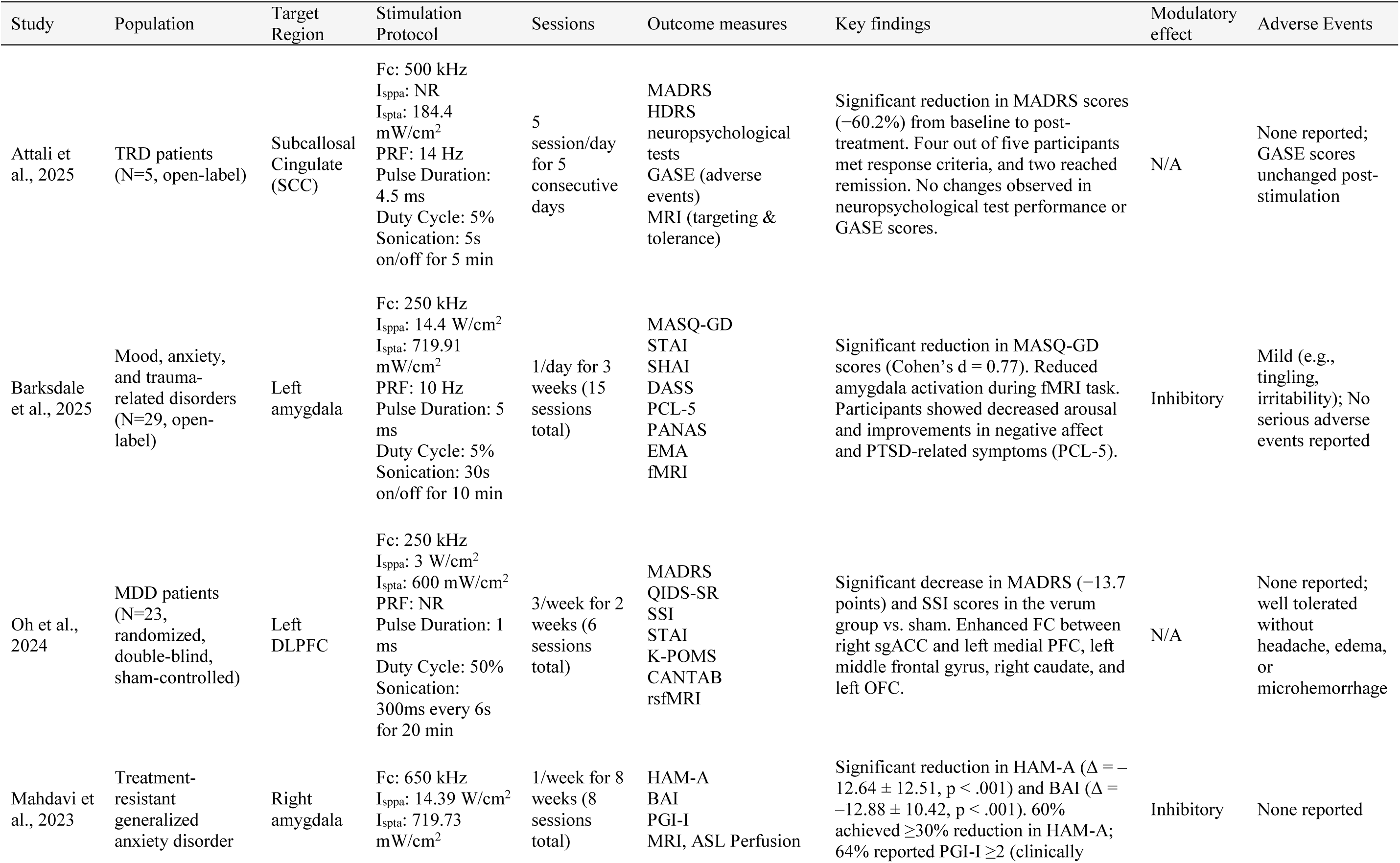
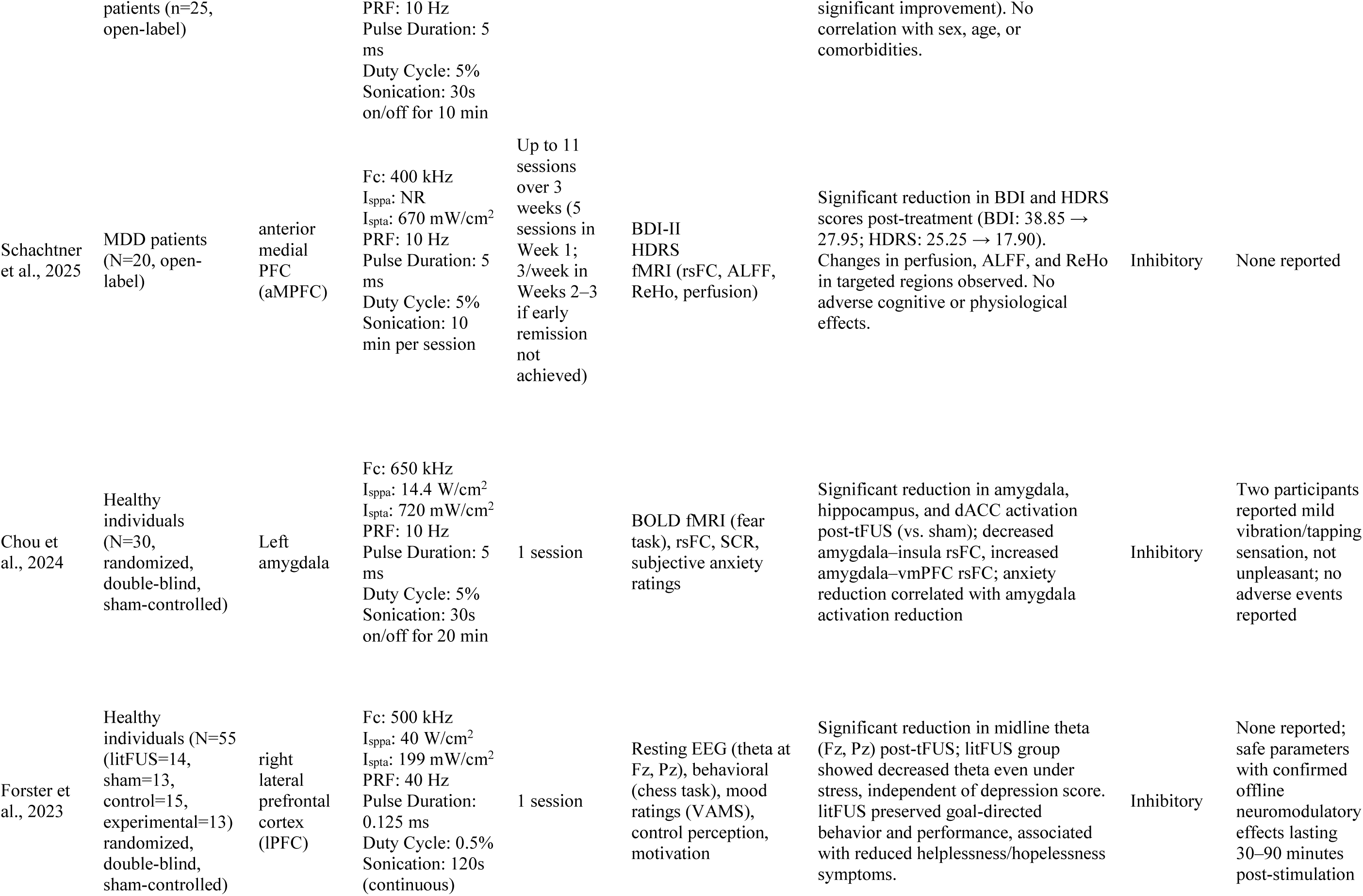
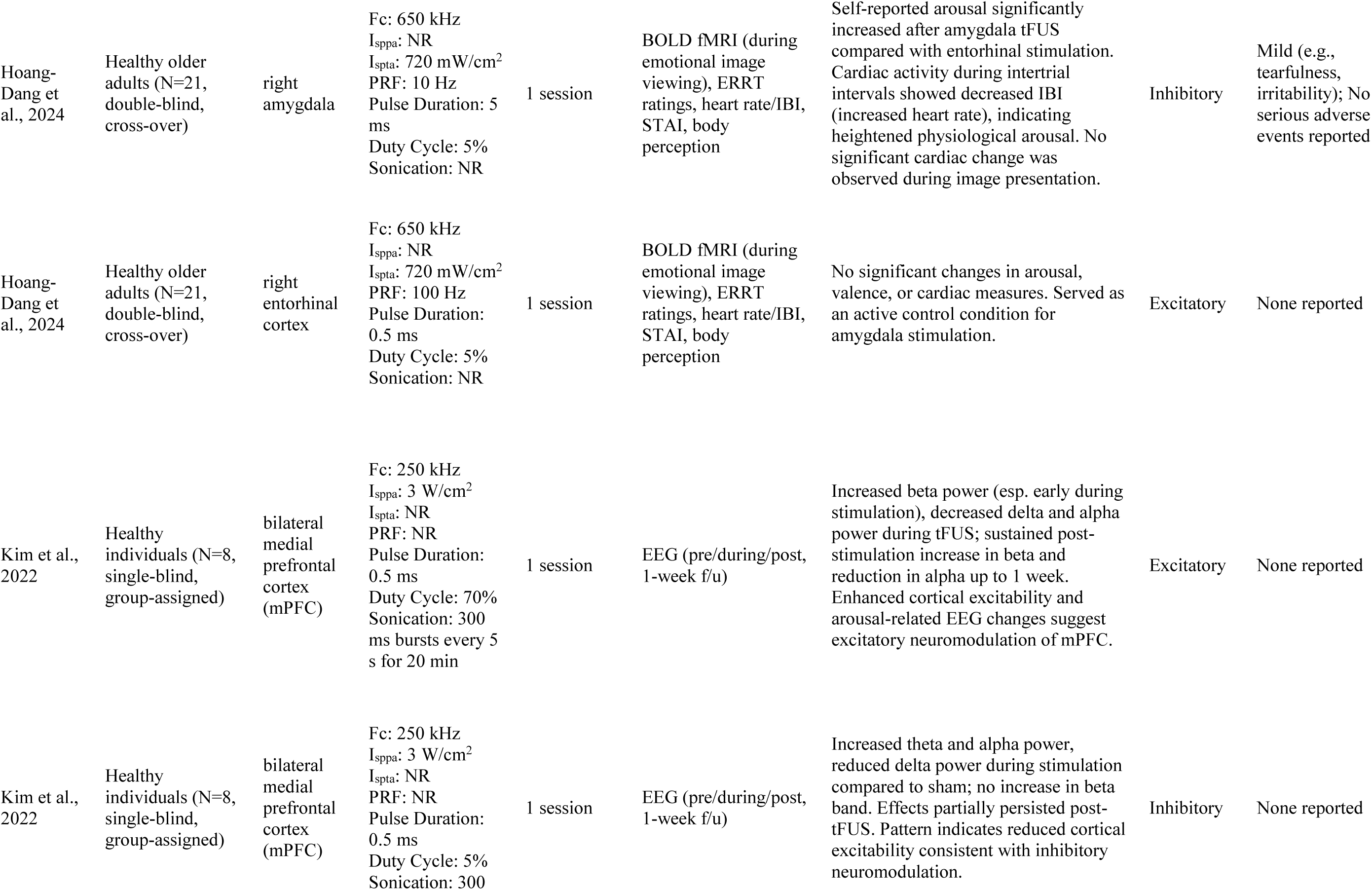
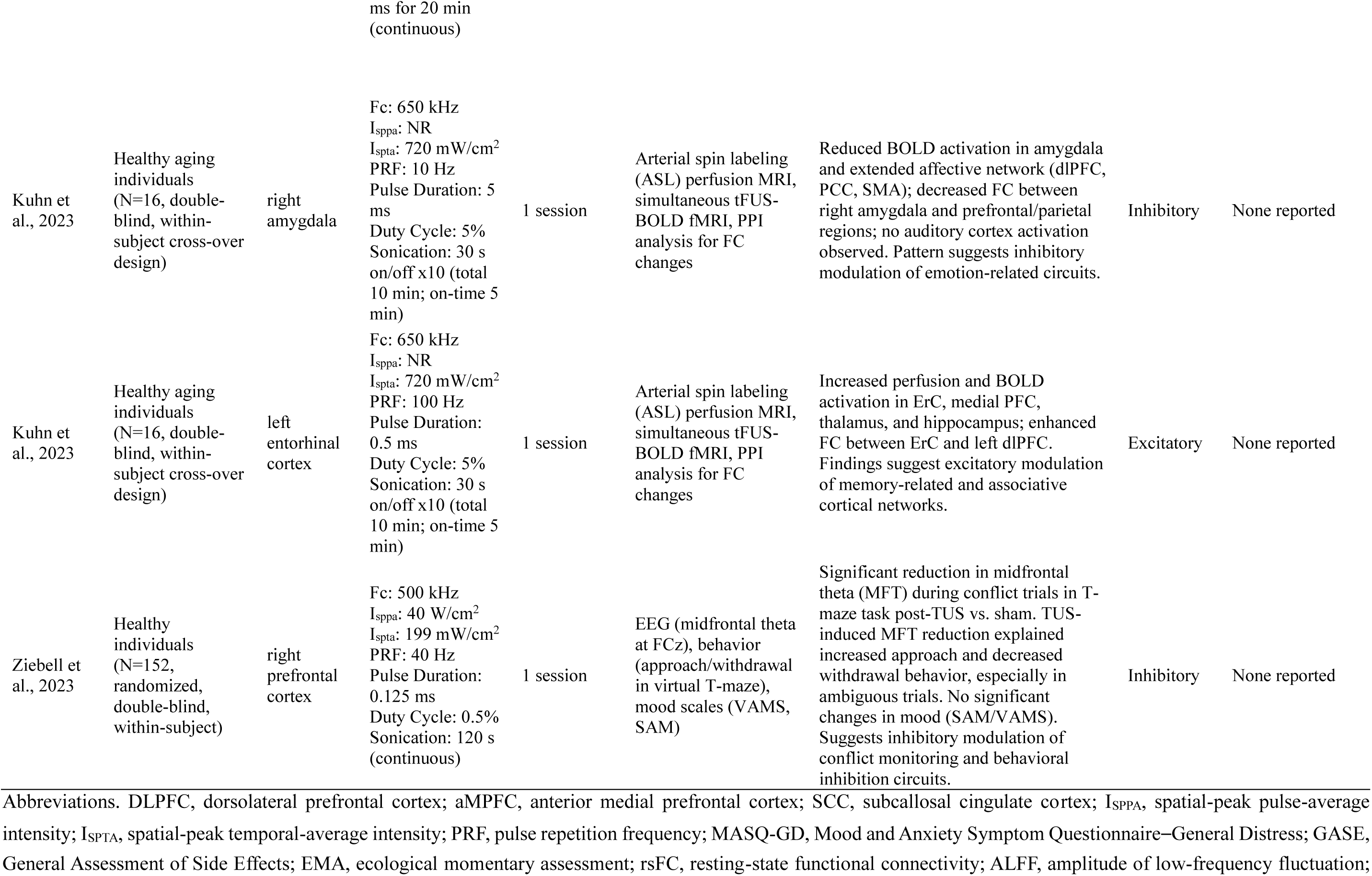
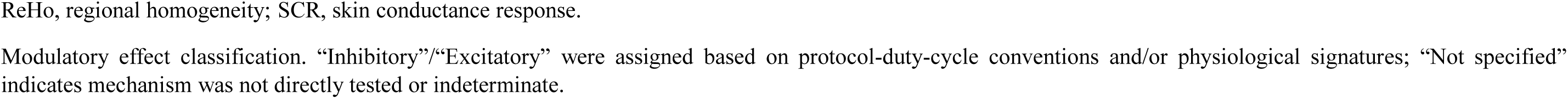
Summary of clinical and experimental tFUS applications for emotion regulation in human studies.

### Amygdala

Five studies in the review employed tFUS targeting the amygdala, comprising one randomized controlled trial (RCT), two double-blind cross-over experiments, and two open-label clinical trials. These studies focused predominantly on its role in modulating anxiety and arousal, with two of these studies (Hoang-Dang et al., 2024; Kuhn et al., 2023) incorporating additional stimulation of the entorhinal cortex within the same participants. Across all studies, the amygdala was selected as a target for its central role in affective reactivity, emotional salience processing, and its dense interconnectivity with limbic-prefrontal networks implicated in anxiety and mood disorders.

tFUS protocols were broadly consistent, using low-intensity stimulation at fundamental frequencies between 250 and 650 kHz, short pulse durations (0.5-5ms), low duty cycles (5%), with pulse repetition frequencies set at 10 Hz. Sonication schedules ranged from single-session to multi-session regimens over several weeks, with unilateral targeting of either the left or right amygdala depending on study design. The majority implemented interleaved on-off blocks of 30 seconds across a 5-to-20-minute period.

The observed neuromodulatory effects were predominantly inhibitory in nature, reporting reductions in amygdala activity and functional connectivity across studies using fMRI, EEG, and arterial spin labeling (ASL). In clinical samples, repeated stimulation of the right amygdala yielded meaningful symptom reductions. Mahdavi et al. (2023) demonstrated significant improvement in anxiety symptomatology, as evidenced by reductions in both Hamilton Anxiety Rating Scale (HAM-A) and Beck Anxiety Inventory (BAI) scores, with 60% of participants achieving a clinically meaningful reduction (≥30%) in HAM-A scores. Similarly, Barksdale et al. (2025) reported significant decreases in negative affect and PTSD-related symptoms following a 15-session tFUS protocol targeting the left amygdala, administered over a three-week period. These effects were accompanied by attenuated amygdala reactivity to emotional stimuli, including reduced BOLD signal during an emotional face-matching task and significant pre-to-post reductions on multiple validated symptom scales. Notably, greater reductions in right amygdala activation were associated with greater improvements in transdiagnostic negative affect, suggesting a potential mechanistic link between subcortical modulation and clinical response.

In nonclinical cohorts, modulation of amygdala was corroborated through converging neuroimaging and physiological evidence. Chou et al. (2024) demonstrated that a single session of tFUS targeting the left amygdala reduced BOLD activation in the amygdala, hippocampus, and dorsal anterior cingulate cortex during a fear-inducing task, with the degree of amygdala signal reduction correlating with decreases in self-reported anxiety. Resting-state fMRI further revealed decreased amygdala-insula connectivity and increased coupling with the ventromedial prefrontal cortex. While changes in skin conductance response were not statistically significant, functional connectivity-based changes suggest a network-level attenuation of arousal.

Complementary findings from another study revealed that right amygdala stimulation increased physiological arousal and subjective emotional reactivity in older adults, indexed by elevated heart rate during intertrial intervals and heightened arousal ratings in response to negative stimuli (Hoang-Dang et al., 2024). These effects were not observed when the entorhinal cortex was targeted in a counterbalanced crossover condition within the same participants, suggesting functional specificity of the amygdala in mediating affective responses to external stimuli.

Kuhn et al. (2023) provided further mechanistic validation of this spatial selectivity using a within-subject crossover design incorporating both BOLD and ASL imaging. Sonication of the right amygdala produced focal increases in perfusion and concurrent decreases in functional connectivity with medial prefrontal and parietal regions. In contrast, stimulation of the entorhinal cortex, administered on a separate day with excitatory parameters, induced increased perfusion and connectivity within memory-related circuits but did not reproduce the amygdala-related effects.

Overall, these findings suggest that amygdala-targeted tFUS predominantly produces inhibitory effects on limbic reactivity and functional connectivity, leading to meaningful symptom reductions in anxiety- and stress-related disorders. However, one study in healthy older adults (Hoang-Dang et al., 2024) reported increased physiological arousal and self-reported reactivity following right amygdala stimulation.

### Prefrontal Cortex

The prefrontal cortex (PFC), a critical hub for top-down regulation of emotion, cognitive control, and behavioral flexibility, is a key target for neuromodulation. Five studies in the review investigated PFC-directed sonication, comprising two randomized, double-blind, sham-controlled trials, two double-blind or single blind experiments, and one open-label clinical trial, targeting dorsolateral, medial, and anterior subregions across both clinical and non-clinical populations (Forster et al., 2023; Kim et al., 2022; Oh et al., 2024; Schachtner et al., 2025; Ziebell et al., 2023). These investigations adopted a range of stimulation protocols, differing in frequency, duty cycle, and session structure, to explore both excitatory and inhibitory neuromodulatory effects.

Sonication parameters spanned fundamental frequencies of 250 to 500 kHz, with pulse durations ranging from 0.125 to 5 ms. Duty cycles varied substantially, from 0.5% in inhibitory paradigms to 70% in excitatory designs. Most studies implemented single-session stimulation, while two clinical trials involved repeated sessions across a two-to three-week period (Oh et al., 2024; Schachtner et al., 2025). Target engagement and neuromodulatory outcomes were assessed using multimodal approaches including EEG, behavioral tasks, clinical symptom scales, and neuroimaging.

In patient populations, prefrontal tFUS was associated with clinically meaningful reductions in depressive symptoms and maladaptive cognitive patterns. A randomized, double-blind, sham-controlled trial applying tFUS to the left dorsolateral prefrontal cortex (DLPFC) across six sessions in individuals with MDD demonstrated significant reductions in depressive severity (−13.7 Montgomery–Åsberg Depression Rating Scale (MADRS) points), suicidal ideation (Suicidal Ideation Questionnaire; SSI), and increased functional connectivity between the subgenual anterior cingulate cortex (sgACC) and key frontostriatal regions, including the medial PFC and orbitofrontal cortex (Oh et al., 2024). Matching these findings, an open-label trial delivering up to 11 sessions of tFUS to the anterior medial PFC (aMPFC), a central node of the default mode network (DMN), reported comparable antidepressant effects, with a 10.9-point reduction on the Beck Depression Inventory-II (BDI-II) and a 4.2-point reduction on the Hamilton Depression Rating Scale (HDRS), alongside significant decreases in repetitive negative thought and improvements in quality of life indices (Schachtner et al., 2025). Sixty percent of participants met standard response criteria, and over one-third achieved remission. No serious adverse events were reported across sessions, and the intervention was well tolerated.

In healthy cohorts, prefrontal tFUS yielded protocol-dependent modulation of cortical excitability and behavior. In a randomized controlled trial, brief low-duty-cycle sonication of the right lateral prefrontal cortex significantly reduced midline theta activity (Fz and Pz), a neural correlate of conflict monitoring and affective distress (Forster et al., 2023). These effects persisted during a learned helplessness paradigm, with tFUS-treated participants maintaining goal-directed behavior and reporting lower helplessness-related affect relative to controls. Building on these findings, a large double-blind within-subject study employing a virtual T-maze task demonstrated that right prefrontal tFUS decreased midfrontal theta (MFT) activity during conflict-inducing decision events, without producing changes in self-reported mood (Ziebell et al., 2023). The tFUS-induced reductions in MFT significantly predicted increased approach and reduced withdrawal behavior, particularly in ambiguous conditions where motivational conflict was highest.

Further evidence of parameter-specific modulation emerged from a within-subject trial that systematically contrasted excitatory and inhibitory sonication to the bilateral medial PFC using a standardized image-guided targeting approach (Kim et al., 2022). Excitatory stimulation delivered using a high-duty-cycle (70%) protocol induced increases in EEG beta power and reductions in alpha and delta bands, with elevated beta activity maintained at follow-up one-week post-stimulation. In contrast, the suppressive protocol (5% duty cycle, continuous bursts) elicited increased theta and alpha activity without beta augmentation, indicative of reduced excitability. These divergent spectral profiles were observed during and after stimulation, and no participants reported adverse effects or auditory confounds.

Collectively, prefrontal tFUS demonstrates consistent antidepressant effects in clinical populations and protocol-dependent modulation of cortical excitability and behavior in healthy cohorts, suggesting its potential as a flexible target for mood regulation.

### Subcallosal Cingulate

A novel application of tFUS to deep brain structures was reported in an open-label pilot trial targeting the subcallosal cingulate (SCC), a region implicated in treatment-resistant depression (TRD) (Attali et al., 2025). Using a personalized, image-guided metalens-based tFUS system, five patients underwent an intensive 5-day (25 sessions) protocol. The intervention produced a significant reduction in depressive symptoms, with a 60.9% mean decrease in MADRS scores by Day 5, and four of the five patients meeting response criteria. No serious adverse events were reported, and follow-up MRI and neurocognitive assessments confirmed the safety and tolerability. Functional connectivity changes were observed between the SCC and the left DLPFC and the right hippocampus.

### Meta-Analytic Findings

A subset of six studies reporting pre- and post-intervention scores on validated affective symptom scales were included in a quantitative synthesis. Hedges’ g values were computed for each study using pooled standard deviations and corrected for small sample bias. The random-effects model yielded a significant overall effect size of g = 0.88, 95% CI [0.47, 1.29], p < .0001, indicating a moderate to large beneficial effect of tFUS across affective domains. Heterogeneity was moderate to high (I^2^ = 64.6%), suggesting variability in effect sizes not attributable to sampling error alone (Table 2, Fig 2).

**Fig 2.**
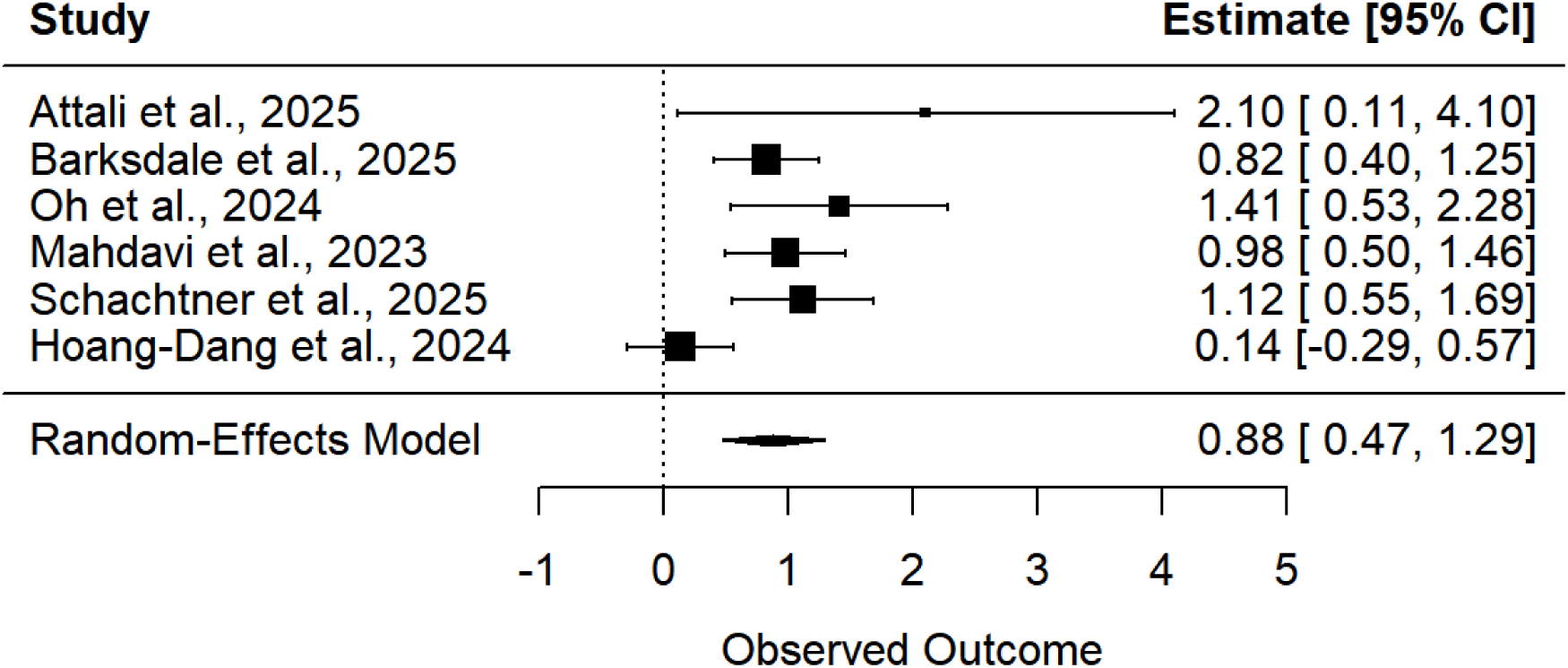
Forest plot of effect sizes (Hedges’ g) for all included studies in the meta-analysis.

**Table 2.** Characteristics and effect sizes of studies included in the meta-analysis.

| Study | N | Target Region | Outcome Measure | Domain | Hedges_g | 95% CI | SE |
| --- | --- | --- | --- | --- | --- | --- | --- |
| Attali et al., 2025 | 5 | Subcallosal Cingulate (SCC) | HDRS | Depression | 2.102053 | [0.11, 4.10] | 1.018 |
| Barksdale et al., 2025 | 29 | Amygdala | MASQ-GD | Anxiety | 0.822986 | [0.40, 1.25] | 0.216 |
| Oh et al., 2024 | 11 | DLPFC | MADRS | Depression | 1.407519 | [0.53, 2.28] | 0.446 |
| Mahdavi et al., 2023 | 25 | Amygdala | HAM-A | Anxiety | 0.977082 | [0.50, 1.46] | 0.246 |
| Schachtner et al., 2025 | 20 | aMPFC | BDI | Depression | 1.117713 | [0.55, 1.69] | 0.290 |
| Hoang-Dang et al., 2024 | 21 | Right amygdala | STAI-T | Anxiety | 0.135407 | [-0.29, 0.57] | 0.219 |

Effect sizes reflect within-subject or pre-post designs, reported as Hedges' g with 95% confidence intervals. SE = standard error. Domain classification was based on the primary clinical target of each study.
N reflects analyzable sample included in meta-analysis, which may differ from total enrolled.
All Hedges' g values computed so that positive values indicate symptom reduction / improvement.

To examine potential differences by symptom domain, subgroup analyses contrasted anxiety- and depression-related outcomes. Although the test for subgroup difference did not reach conventional significance (p = 0.0907), domain accounted for 31.7% of between-study heterogeneity (R^2^), and the pooled effect size was larger for depression (g = 1.31) than for anxiety (g = 0.67), although this difference did not reach statistical significance (Fig 3). Sensitivity analyses excluding ultra-small samples (N < 10; e.g., Attali et al., 2025) yielded a comparable pooled effect (g = 0.81, 95% CI [0.23, 1.39], I^2^ = 0%), indicating that the overall findings were not driven by very small trials. Publication bias was not formally assessed with Egger’s test due to the limited number of studies (k < 10), which would render the test underpowered.

**Fig 3.**
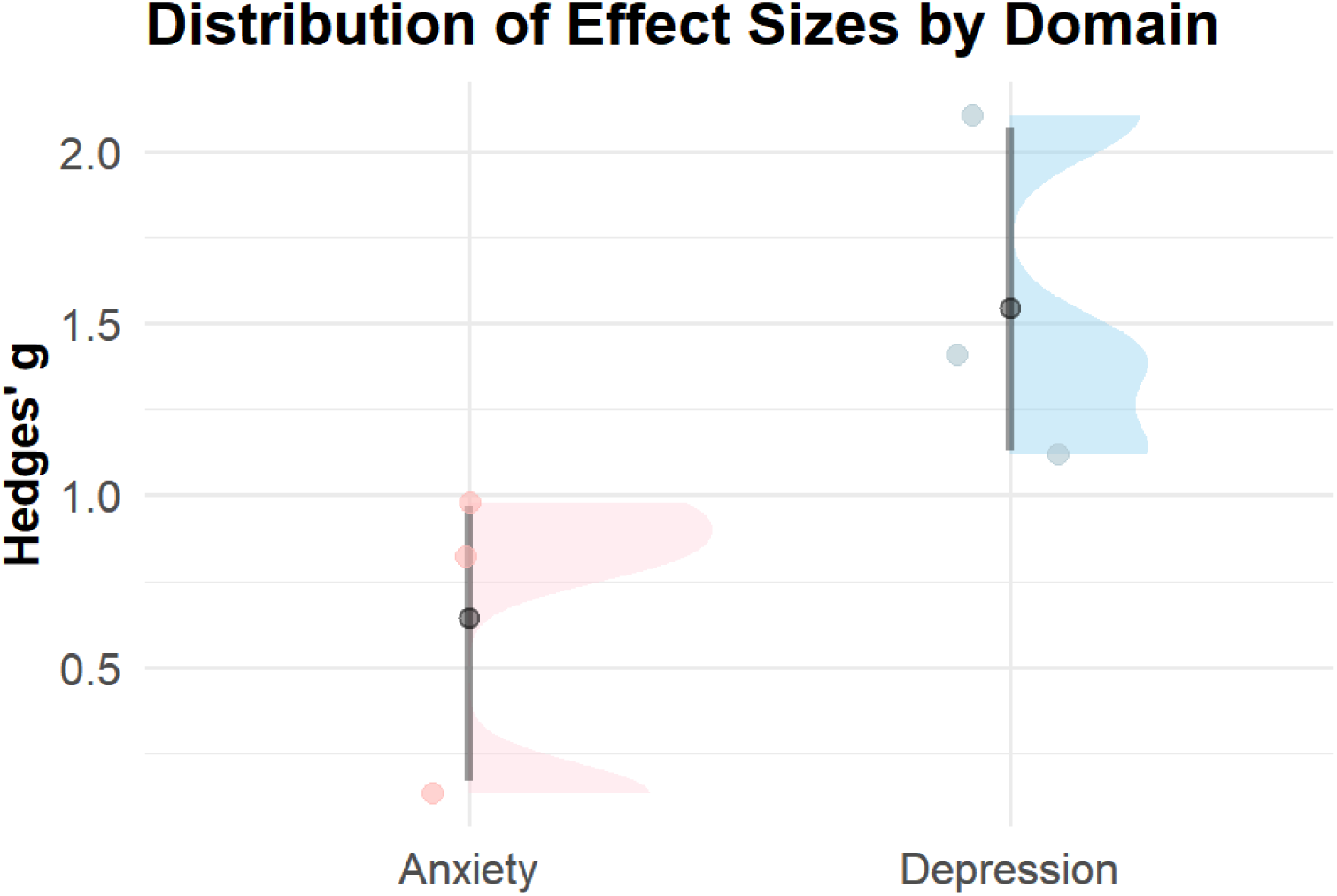
Distribution of effect sizes by clinical domain (Anxiety vs. Depression)

## Discussion

The present systematic review provides the first comprehensive synthesis of human studies employing tFUS with the explicit aim of modulating emotion regulation-related affective outcomes. The findings suggest that tFUS is a promising neuromodulatory approach capable of engaging core emotion regulation networks, including the amygdala, prefrontal cortex (PFC), and subcallosal cingulate cortex (SCC), with measurable impact on mood, anxiety, and physiological arousal. Across 11 included studies, tFUS consistently modulated neural circuits implicated in emotion regulation and yielded clinically relevant symptom change, with a significant pooled effect size (Hedges’ g = 0.88), suggesting moderate-to-large effects.

Increasing preclinical and translational evidence further supports LIFU as a next-generation neuromodulation modality with unique advantages for circuit-specific intervention (Badran & Peng, 2024; Fini & Tyler, 2017). Distinct from transcranial magnetic stimulation (TMS) or transcranial direct current stimulation (tDCS), which are constrained by limited depth penetration and diffuse current spread (Aderinto et al., 2024; Razza et al., 2023), tFUS offers submillimeter spatial precision and the ability to reach deep limbic and paralimbic targets implicated in affective dysregulation, thereby enabling precise engagement of emotion regulation circuits. This capability is particularly relevant for mood and anxiety disorders, which involve distributed dysregulation across corticolimbic networks, including medial prefrontal, anterior cingulate, and limbic structures (Pizzagalli & Roberts, 2022). Moreover, tFUS is well tolerated across both single-session and repeated-session protocols, with no serious adverse events reported in the reviewed studies in line with systematic reviews of tFUS safety (Sarica et al., 2022).

Subgroup analyses revealed domain-specific trends, with larger effects observed in studies targeting depressive symptoms (g = 1.31) compared to those addressing anxiety (g = 0.67). Although the subgroup difference did not reach statistical significance, the difference accounted for over 30% of between-study heterogeneity and may reflect systematic variation in target selection, stimulation protocol, and outcome metrics. Depression-focused protocols more frequently employed multi-session stimulation of the dorsolateral or medial PFC, regions implicated in mood regulation and executive control, while anxiety-focused studies predominantly targeted the amygdala, a core hub for salience detection and emotional reactivity within the emotion regulation network (Ashworth et al., 2021; Gou et al., 2023; Kan et al., 2023).

From a clinical perspective, the accumulating evidence also infers that tFUS is a candidate intervention for treatment-resistant populations. In trials involving patients with major depressive disorder unresponsive to conventional pharmacotherapy, repeated prefrontal or SCC-directed tFUS was associated with rapid and clinically meaningful symptom reductions, with no reported serious adverse events (Attali et al., 2025; Oh et al., 2024; Schachtner et al., 2025). Compared with other circuit-targeted interventions, such as deep brain stimulation (DBS) or high-intensity MR-guided focused ultrasound, LIFU provides a non-invasive alternative that can reach cortical and subcortical nodes with a favorable safety profile. Notably, the magnitude of improvement observed in our synthesis (g = 0.88 overall; g = 1.31 for depression) is broadly comparable to the clinical benefits reported for TMS in large trials such as BRIGhTMIND (Morriss et al., 2024), although direct comparisons are limited by differences in study design, sample size, and follow-up duration.

Translation of tFUS into routine clinical practice faces several practical challenges. MR-guided platforms offer highly accurate targeting of cortical and deep brain structures but demand specialized facilities, longer setup times, and higher operational cost, whereas non-MR-guided neuronavigation systems are more accessible and cost-efficient but may offer less accuracy for deep targets (Legon et al., 2018; Verhagen et al., 2019). Widespread adoption will require standardized treatment protocols, specialized operator training, and strategies to optimize adherence during multi-session interventions, particularly in psychiatric contexts where maintenance neuromodulation may be indicated.

Rather than serving solely as a standalone intervention, tFUS may ultimately be most effective as part of an integrated, multimodal intervention framework (Davidson et al., 2025). Its capacity to induce neuroplasticity and modulate network connectivity creates opportunities for synergy with psychotherapy, for example by enhancing emotional regulation and cognitive restructuring during behavioral interventions (Saccenti et al., 2024; Vanderhasselt et al., 2021). Similarly, in pharmacotherapy, tFUS may augment drug effects by directly engaging neural circuits that mediate therapeutic response, thereby improving outcomes in partial or non-responders (Tao et al., 2024; Wang et al., 2021).

Despite promising findings, several caveats warrant caution in interpretation. First, only a small subset of included trials employed randomized, sham-controlled designs, and most were open-label or pilot in nature. Thus, while pooled effect sizes were significant, the certainty of evidence remains low, and efficacy cannot yet be established with confidence. Second, few studies reported pre-registered protocols, a priori sample size estimations, or comprehensive bias-mitigation procedures, all of which are critical for reproducibility and reliability. Risk-of-bias assessments further highlighted concerns regarding randomization procedures, blinding, and outcome reporting in several trials. Third, although safety profiles were favorable in short-term studies, the durability of effects and long-term safety in more severe or treatment-resistant clinical populations remain unknown. Larger sham-controlled RCTs with prolonged follow-up are required to address these gaps.

For tFUS to fulfill its therapeutic promise in emotion regulation, both the scientific foundation and the clinical infrastructure must be strategically advanced. Future investigations should prioritize large-scale randomized controlled trials with harmonized stimulation protocols, validated and domain-sensitive outcome measures, and long-term follow-up to determine efficacy, durability, and safety across diverse populations. Mechanistic studies integrating high-resolution neuroimaging, electrophysiological markers, and behavioral assessments will be essential for identifying robust biomarkers of response and refining target selection. In parallel, technical advances toward portable, cost-effective, and user-friendly tFUS systems, coupled with standardized training frameworks, will be critical for enabling widespread clinical adoption. Through these converging efforts, tFUS has the potential to mature from an experimental neuromodulation tool into a versatile, precision-based intervention capable of delivering sustained improvements in emotion regulation and mental health outcomes.

## Data Availability

All data generated or analyzed in this study are from previously published articles and are summarized within the manuscript and supplementary files. Extracted datasets used for the meta-analysis are available from the corresponding author upon reasonable request.

## Acknowledgments

Not applicable.

## References

Abend, R., Sar-El, R., Gonen, T., Jalon, I., Vaisvaser, S., Bar-Haim, Y., & Hendler, T. (2019). Modulating emotional experience using electrical stimulation of the medial-prefrontal cortex: a preliminary tDCS-fMRI study. Neuromodulation: Technology at the Neural Interface, 22(8), 884–893.

Aderinto, N., Olatunji, G., Muili, A. et al. A narrative review of non-invasive brain stimulation techniques in neuropsychiatric disorders: current applications and future directions. Egypt J Neurol Psychiatry Neurosurg 60, 50 (2024). 10.1186/s41983-024-00824-w

Andò, A., Vasilotta, M. L., & Zennaro, A. (2021). The modulation of emotional awareness using non-invasive brain stimulation techniques: A literature review on TMS and tDCS. Journal of Cognitive Psychology, 33(8), 993–1010.

Ashworth, E., Brooks, S. J., & Schiöth, H. B. (2021). Neural activation of anxiety and depression in children and young people: A systematic meta-analysis of fMRI studies. Psychiatry Research: Neuroimaging, 311, 111272. 10.1016/j.pscychresns.2021.111272

Atkinson-Clement, C., Junor, A., & Kaiser, M. (2025). Neuromodulation perception by the general public. Scientific Reports, 15(1), 5584. 10.1038/s41598-025-89437-8

Attali, D., Tiennot, T., Manuel, T. J., Daniel, M., Houdouin, A., Annic, P., Dizeux, A., Haroche, A., Dadi, G., Henensal, A., Moyal, M., Le Berre, A., Paolillo, C., Charron, S., Debacker, C., Lui, M., Lekcir, S., Mancusi, R., Gallarda, T.,…Plaze, M. (2025). Deep transcranial ultrasound stimulation using personalized acoustic metamaterials improves treatment-resistant depression in humans. Brain Stimul, 18(3), 1004–1014. 10.1016/j.brs.2025.04.018

Badran, B. W., & Peng, X. (2024). Transcranial focused ultrasound (tFUS): a promising noninvasive deep brain stimulation approach for pain. In: Springer International Publishing Cham.

Barksdale, B. R., Enten, L., DeMarco, A., Kline, R., Doss, M. K., Nemeroff, C. B., & Fonzo, G. A. (2025). Low-intensity transcranial focused ultrasound amygdala neuromodulation: a double-blind sham-controlled target engagement study and unblinded single-arm clinical trial. Mol Psychiatry. 10.1038/s41380-025-03033-w

Bertocci, M., Chase, H., Graur, S., Stiffler, R., Edmiston, E., Coffman, B., Greenberg, B., & Phillips, M. (2021). The impact of targeted cathodal transcranial direct current stimulation on reward circuitry and affect in Bipolar Disorder. Molecular psychiatry, 26(8), 4137–4145.

Caffaratti, H., Slater, B., Shaheen, N., Rhone, A., Calmus, R., Kritikos, M., Kumar, S., Dlouhy, B., Oya, H., Griffiths, T., Boes, A. D., Trapp, N., Kaiser, M., Sallet, J., Banks, M. I., Howard, M. A., Zanaty, M., & Petkov, C. I. (2025). Neuromodulation with Ultrasound: Hypotheses on the Directionality of Effects and Community Resource. In: eLife Sciences Publications, Ltd.

Chou, T., Deckersbach, T., Guerin, B., Sretavan Wong, K., Borron, B. M., Kanabar, A., Hayden, A. N., Long, M. P., Daneshzand, M., Pace-Schott, E. F., & Dougherty, D. D. (2024). Transcranial focused ultrasound of the amygdala modulates fear network activation and connectivity. Brain Stimul, 17(2), 312–320. 10.1016/j.brs.2024.03.004

Cisler, J. M., & Olatunji, B. O. (2012). Emotion regulation and anxiety disorders. Current psychiatry reports, 14(3), 182–187.

Davidson, B., Xhima, K., Cosgrove, R., Hamani, C., Eitan, R., Rezai, A., LeBlang, S., Philip, N. S., & Lipsman, N. (2025). A roadmap for focused ultrasound applications in psychiatry: Proceedings of the 2024 symposium on focused ultrasound in psychiatry (FUS-PULSE). Brain stimulation, 18(5), 1651–1662. 10.1016/j.brs.2025.08.012

Eres, R., Lim, M. H., Lanham, S., Jillard, C., & Bates, G. (2021). Loneliness and emotion regulation: Implications of having social anxiety disorder. Australian Journal of Psychology, 73(1), 46–56.

Fini, M., & Tyler, W. J. (2017). Transcranial focused ultrasound: a new tool for non-invasive neuromodulation. International Review of Psychiatry, 29(2), 168–177.

Forster, A., Rodrigues, J., Ziebell, P., Sanguinetti, J. L., Allen, J. J. B., & Hewig, J. (2023). Transcranial focused ultrasound modulates the emergence of learned helplessness via midline theta modification. J Affect Disord, 329, 273–284. 10.1016/j.jad.2023.02.032

Gou, X.-y., Li, Y.-x., Guo, L.-x., Zhao, J., Zhong, D.-l., Liu, X.-b., Xia, H.-s., Fan, J., Zhang, Y., Ai, S.-c., Huang, J.-x., Li, H.-r., Li, J., & Jin, R.-j. (2023). The conscious processing of emotion in depression disorder: a meta-analysis of neuroimaging studies [Systematic Review]. Frontiers in Psychiatry, Volume 14 - 2023. 10.3389/fpsyt.2023.1099426

Gross, J. J. (2015). Emotion regulation: Current status and future prospects. Psychological inquiry, 26(1), 1–26.

Guidetti, C., Feeney, A., Hock, R. S., Iovieno, N., Ortiz, J. M. H., Fava, M., & Papakostas, G. I. (2025). Antidepressants in the acute treatment of post-traumatic stress disorder in adults: a systematic review and meta-analysis. International Clinical Psychopharmacology, 40(3), 138–147.

Hamilton, M. (1959). Hamilton anxiety rating scale (HAM-A). J Med, 61(4), 81–8.

Hedges, L. V., & Olkin, I. (2014). Statistical methods for meta-analysis. Academic press.

Hoang-Dang, B., Halavi, S. E., Rotstein, N. M., Spivak, N. M., Dang, N. H., Cvijanovic, L., Hiller, S. H., Vallejo-Martelo, M., Rosenberg, B. M., Swenson, A., Becerra, S., Sun, M., Revett, M. E., Kronemyer, D., Berlow, R., Craske, M. G., Suthana, N., Monti, M. M., Zbozinek, T. D.,…Kuhn, T. P. (2024). Transcranial Focused Ultrasound Targeting the Amygdala May Increase Psychophysiological and Subjective Negative Emotional Reactivity in Healthy Older Adults. Biol Psychiatry Glob Open Sci, 4(5), 100342. 10.1016/j.bpsgos.2024.100342

Jin, J., Pei, G., Ji, Z., Liu, X., Yan, T., Li, W., & Suo, D. (2024). Transcranial focused ultrasound precise neuromodulation: a review of focal size regulation, treatment efficiency and mechanisms. Frontiers in Neuroscience, 18, 1463038.

Kan, R. L. D., Padberg, F., Giron, C. G., Lin, T. T. Z., Zhang, B. B. B., Brunoni, A. R., & Kranz, G. S. (2023). Effects of repetitive transcranial magnetic stimulation of the left dorsolateral prefrontal cortex on symptom domains in neuropsychiatric disorders: a systematic review and cross-diagnostic meta-analysis. The Lancet Psychiatry, 10(4), 252–259. 10.1016/S2215-0366(23)00026-3

Kang, J., Lee, H., Yu, S. et al. Effects and safety of transcranial direct current stimulation on multiple health outcomes: an umbrella review of randomized clinical trials. Mol Psychiatry 29, 3789– 3801 (2024). 10.1038/s41380-024-02624-3

Kim, Y. G., Kim, S. E., Lee, J., Hwang, S., Yoo, S.-S., & Lee, H. W. (2022). Neuromodulation using transcranial focused ultrasound on the bilateral medial prefrontal cortex. Journal of clinical medicine, 11(13), 3809.

Kuhn, T., Spivak, N. M., Dang, B. H., Becerra, S., Halavi, S. E., Rotstein, N., Rosenberg, B. M., Hiller, S., Swenson, A., Cvijanovic, L., Dang, N., Sun, M., Kronemyer, D., Berlow, R., Revett, M. R., Suthana, N., Monti, M. M., & Bookheimer, S. (2023). Transcranial focused ultrasound selectively increases perfusion and modulates functional connectivity of deep brain regions in humans. Front Neural Circuits, 17, 1120410. 10.3389/fncir.2023.1120410

Legon, W., Ai, L., Bansal, P., & Mueller, J. K. (2018). Neuromodulation with single-element transcranial focused ultrasound in human thalamus. Hum Brain Mapp, 39(5), 1995–2006. 10.1002/hbm.23981

Lincoln, T. M., Schulze, L., & Renneberg, B. (2022). The role of emotion regulation in the characterization, development and treatment of psychopathology. Nature Reviews Psychology, 1(5), 272–286.

Liu, W., Leng, Y. S., Zou, X. H., Cheng, Z. Q., Yang, W., & Li, B. J. (2017). Affective processing in non-invasive brain stimulation over prefrontal cortex. Frontiers in Human Neuroscience, 11, 439.

Mahdavi, K. D., Jordan, S. E., Jordan, K. G., Rindner, E. S., Haroon, J. M., Habelhah, B., Becerra, S. A., Surya, J. R., Venkatraman, V., Zielinski, M. A., Spivak, N. M., Bystritsky, A., & Kuhn, T. P. (2023). A pilot study of low-intensity focused ultrasound for treatment-resistant generalized anxiety disorder. J Psychiatr Res, 168, 125–132. 10.1016/j.jpsychires.2023.10.039

McLean, C. P., & Foa, E. B. (2017). Emotions and emotion regulation in posttraumatic stress disorder. Current opinion in psychology, 14, 72–77.

Moher, D., Liberati, A., Tetzlaff, J., Altman, D. G., & PRISMA Group*, t. (2009). Preferred reporting items for systematic reviews and meta-analyses: the PRISMA statement. Annals of internal medicine, 151(4), 264–269.

Montgomery, S. A., & Åsberg, M. (1979). A new depression scale designed to be sensitive to change. The British journal of psychiatry, 134(4), 382–389.

Morriss, R., Briley, P. M., Webster, L., Abdelghani, M., Barber, S., Bates, P., Brookes, C., Hall, B., Ingram, L., Kurkar, M., Lankappa, S., Liddle, P. F., McAllister-Williams, R. H., O’Neil-Kerr, A., Pszczolkowski, S., Suazo Di Paola, A., Walters, Y., & Auer, D. P. (2024). Connectivity-guided intermittent theta burst versus repetitive transcranial magnetic stimulation for treatment-resistant depression: a randomized controlled trial. Nature Medicine, 30(2), 403–413. 10.1038/s41591-023-02764-z

Nierenberg, A. A., Petersen, T. J., & Alpert, J. E. (2003). Prevention of relapse and recurrence in depression: the role of long-term pharmacotherapy and psychotherapy. Journal of Clinical Psychiatry, 64(15), 13–17.

Oathes, D. J., Balderston, N. L., Kording, K. P., DeLuisi, J. A., Perez, G. M., Medaglia, J. D., Fan, Y., Duprat, R. J., Satterthwaite, T. D., & Sheline, Y. I. (2021). Combining transcranial magnetic stimulation with functional magnetic resonance imaging for probing and modulating neural circuits relevant to affective disorders. Wiley Interdisciplinary Reviews: Cognitive Science, 12(4), e1553.

Oh, J., Ryu, J. S., Kim, J., Kim, S., Jeong, H. S., Kim, K. R., Kim, H. C., Yoo, S. S., & Seok, J. H. (2024). Effect of Low-Intensity Transcranial Focused Ultrasound Stimulation in Patients With Major Depressive Disorder: A Randomized, Double-Blind, Sham-Controlled Clinical Trial. Psychiatry Investig, 21(8), 885–896. 10.30773/pi.2024.0016

Page, M. J., McKenzie, J. E., Bossuyt, P. M., Boutron, I., Hoffmann, T. C., Mulrow, C. D., Shamseer, L., Tetzlaff, J. M., Akl, E. A., Brennan, S. E., Chou, R., Glanville, J., Grimshaw, J. M., Hróbjartsson, A., Lalu, M. M., Li, T., Loder, E. W., Mayo-Wilson, E., McDonald, S., McGuinness, L. A., … Moher, D. (2021). The PRISMA 2020 statement: an updated guideline for reporting systematic reviews. BMJ (Clinical research ed.), 372, n71. 10.1136/bmj.n71

Pizzagalli, D. A., & Roberts, A. C. (2022). Prefrontal cortex and depression. Neuropsychopharmacology, 47(1), 225–246. 10.1038/s41386-021-01101-7

Razza, L. B., Luethi, M. S., Zanão, T., De Smet, S., Buchpiguel, C., Busatto, G., Pereira, J., Klein, I., Kappen, M., Moreno, M., Baeken, C., Vanderhasselt, M.-A., & Brunoni, A. R. (2023). Transcranial direct current stimulation versus intermittent theta-burst stimulation for the improvement of working memory performance. International Journal of Clinical and Health Psychology, 23(1), 100334. 10.1016/j.ijchp.2022.100334

Regenold, W.T., Deng, ZD. & Lisanby, S.H. Noninvasive neuromodulation of the prefrontal cortex in mental health disorders. Neuropsychopharmacol. 47, 361–372 (2022). 10.1038/s41386-021-01094-3

Sabé, M., Hyde, J., Cramer, C., Eberhard, A.-L., Crippa, A., Brunoni, A. R., Aleman, A., Kaiser, S., Baldwin, D. S., & Garner, M. (2024). Transcranial magnetic stimulation and transcranial direct current stimulation across mental disorders: a systematic review and dose-response meta-analysis. JAMA network Open, 7(5), e2412616–e2412616.

Saccenti, D., Lauro, L. J. R., Crespi, S. A., Moro, A. S., Vergallito, A., Grgič, R. G., Pretti, N., Lamanna, J., & Ferro, M. (2024). Boosting Psychotherapy With Noninvasive Brain Stimulation: The Whys and Wherefores of Modulating Neural Plasticity to Promote Therapeutic Change. Neural Plasticity, 2024(1), 7853199. 10.1155/np/7853199

Sanguinetti, J. L., Hameroff, S., Smith, E. E., Sato, T., Daft, C. M., Tyler, W. J., & Allen, J. J. (2020). Transcranial focused ultrasound to the right prefrontal cortex improves mood and alters functional connectivity in humans. Frontiers in Human Neuroscience, 14, 52.

Sarica, C., Nankoo, J.-F., Fomenko, A., Grippe, T. C., Yamamoto, K., Samuel, N., Milano, V., Vetkas, A., Darmani, G., Cizmeci, M. N., Lozano, A. M., & Chen, R. (2022). Human Studies of Transcranial Ultrasound neuromodulation: A systematic review of effectiveness and safety. Brain stimulation, 15(3), 737–746. 10.1016/j.brs.2022.05.002

Schachtner, J. N., Dahill-Fuchel, J. F., Allen, K. E., Bawiec, C. R., Hollender, P. J., Ornellas, S. B., Konecky, S. D., Achrol, A. S., & Allen, J. J. B. (2025). Transcranial focused ultrasound targeting the default mode network for the treatment of depression. Front Psychiatry, 16, 1451828. 10.3389/fpsyt.2025.1451828

Sharif F, Harmer CJ, Klein-Flügge MC, Tan H. Novel NIBS in psychiatry: Unveiling TUS and TI for research and treatment. Brain and Neuroscience Advances. 2025;9. doi:10.1177/23982128251322241

Spielberger, C. D., Gonzalez-Reigosa, F., Martinez-Urrutia, A., Natalicio, L. F., & Natalicio, D. S. (1971). The state-trait anxiety inventory. Revista Interamericana de Psicologia/Interamerican journal of psychology, 5(3 & 4).

Tao, Y., Liang, Q., Zhang, F., Guo, S., Fan, L., & Zhao, F. (2024). Efficacy of non-invasive brain stimulation combined with antidepressant medications for depression: a systematic review and meta-analysis of randomized controlled trials. Systematic Reviews, 13(1), 92. 10.1186/s13643-024-02480-w

Tanaka, M., He, Z., Han, S., & Battaglia, S. (2025). Noninvasive brain stimulation: a promising approach to study and improve emotion regulation. In (Vol. 19, pp. 1633936): Frontiers Media SA.

Toth, J., Kurtin, D. L., Brosnan, M., & Arvaneh, M. (2024). Opportunities and obstacles in non-invasive brain stimulation [Perspective]. Frontiers in Human Neuroscience, Volume 18–2024. 10.3389/fnhum.2024.1385427

Vanderhasselt, M.-A., Dedoncker, J., De Raedt, R., & Baeken, C. (2021). Combination of tDCS with Psychotherapy and Neurobehavioral Interventions: Systematic Review and Mechanistic Principles for Future Clinical Trials. In A. R. Brunoni, M. A. Nitsche, & C. K. Loo (Eds.), Transcranial Direct Current Stimulation in Neuropsychiatric Disorders: Clinical Principles and Management (pp. 741–755). Springer International Publishing. 10.1007/978-3-030-76136-3_39

Verhagen, L., Gallea, C., Folloni, D., Constans, C., Jensen, D. E., Ahnine, H., Roumazeilles, L., Santin, M., Ahmed, B., Lehericy, S., Klein-Flügge, M. C., Krug, K., Mars, R. B., Rushworth, M. F., Pouget, P., Aubry, J. F., & Sallet, J. (2019). Offline impact of transcranial focused ultrasound on cortical activation in primates. Elife, 8. 10.7554/eLife.40541

Villalobos, D., Pacios, J., & Vázquez, C. (2021). Cognitive control, cognitive biases and emotion regulation in depression: A new proposal for an integrative interplay model. Frontiers in Psychology, 12, 628416.

Wang, J., Luo, H., Schülke, R., Geng, X., Sahakian, B. J., & Wang, S. (2021). Is transcranial direct current stimulation, alone or in combination with antidepressant medications or psychotherapies, effective in treating major depressive disorder? A systematic review and meta-analysis. BMC Medicine, 19(1), 319. 10.1186/s12916-021-02181-4

Zhang, T., Pan, N., Wang, Y., Liu, C., & Hu, S. (2021). Transcranial focused ultrasound neuromodulation: a review of the excitatory and inhibitory effects on brain activity in human and animals. Frontiers in Human Neuroscience, 15, 749162.

Zhou, D., Li, X., Wei, S., Yu, C., Wang, D., Li, Y., Li, J., Liu, J., Li, S., & Zhuang, W. (2024). Transcranial direct current stimulation combined with repetitive transcranial magnetic stimulation for depression: a randomized clinical trial. JAMA network Open, 7(11), e2444306–e2444306.

Ziebell, P., Rodrigues, J., Forster, A., Sanguinetti, J. L., Allen, J. J., & Hewig, J. (2023). Inhibition of midfrontal theta with transcranial ultrasound explains greater approach versus withdrawal behavior in humans. Brain Stimul, 16(5), 1278–1288. 10.1016/j.brs.2023.08.011

