## Supplementary Table 1 for "Transcranial Focused Ultrasound for Emotion Regulation: A Systematic Review and Quantitative Summary of Human Studies"

**Supplementary Table 1.** Risk of bias assessment in included studies

| Study | Randomization/ Blinding | Control adequacy | Outcome validity | Reporting completeness | Follow-up/ Analysis consistency | Overall |
|---|---|---|---|---|---|---|
| Attali et al., 2025 | High | High | Moderate | Low | Moderate | High |
| Barksdale et al., 2025 | Mixed | Moderate | Low | Low | Moderate | Moderate |
| Oh et al., 2024 | Low | Low | Low | Low | Moderate | Low |
| Mahdavi et al., 2023 | High | High | Moderate | Moderate | High | High |
| Schachtner et al., 2025 | Low | Low | Low | Low | Low | Low |
| Chou et al., 2024 | Low | Low | Low | Low | Low | Low |
| Forster et al., 2023 | Low | Low | Low | Low | Low | Low |
| Hoang-Dang et al., 2024 | Low | Moderate | Low | Low | Moderate | Moderate |
| Kim et al., 2022 | Moderate | Low | Low | Low | Moderate | Moderate |
| Kuhn et al., 2023 | Low | Moderate | Low | Low | Moderate | Moderate |
| Ziebell et al., 2023 | Low | Low | Low | Low | Moderate | Low |

1. Randomization blinding: clarity of participant allocation methods and whether participants, investigators, or outcome assessors were blinded.
2. Control adequacy: appropriateness of comparator (e.g., sham, baseline, active control for minimizing bias).
3. Outcome validity: use of validated and reliable clinical, behavioral, or neuroimaging outcome measures.
4. Reporting completeness: transparency in reporting methods, results, and adverse events.
5. Follow-up/Analysis consistency: adequacy of follow-up procedures and statistical analyses, including handling of missing data.
