## Supplementary 1 for "Transcranial Focused Ultrasound for Emotion Regulation: A Systematic Review and Quantitative Summary of Human Studies"

**Supplementary 1.** Information Sources and Search Strategy

<u>Databases and Scope:</u>

A systematic literature search was conducted across three electronic databases: PubMed/Medline, Embase and PsychINFO. The last search was performed on June 24, 2025. No restrictions were placed on publication year. Only studies published in English and conducted in humans were included.

Reference lists of included studies and relevant reviews were manually screened to identify additional eligible records. Duplicate citations were removed using EndNote 21, followed by manual verification prior to screening.

<u>Search Keywords and Strategy:</u>

The same conceptual search string was applied across databases, with minor syntax adjustments to match each platform's field tags and subject headings.

(("transcranial focused ultrasound"[Title/Abstract] OR "focused ultrasound neuromodulation"[Title/Abstract] OR "low-intensity focused ultrasound"[Title/Abstract] OR LIFU[Title/Abstract] OR tFUS[Title/Abstract]) AND ("emotion regulation"[Title/Abstract] OR emotion*[Title/Abstract] OR affect*[Title/Abstract] OR anxiety[Title/Abstract] OR depressive[Title/Abstract] OR depression[Title/Abstract] OR PTSD[Title/Abstract] OR "post-traumatic stress"[Title/Abstract] OR stress[Title/Abstract])) AND (humans[MeSH Terms]) AND (english[Language])
